# A LITERATURE REVIEW ON COMPLEMENTARY THERAPIES IN AUTISM CHILDREN: A NURSING SCIENCE PHILOSOPHY

**DOI:** 10.1101/2022.09.21.22280230

**Authors:** Dera Alfiyanti, Moses Glorino Rumambo Pandin, Imelda Rizky Rahayuningtyas

## Abstract

**Introduction:** Autism is a neurodevelopmental disorder that can cause repetitive behavior, limited activity, and a lack of ability to communicate and socialize. The purpose of this *literature review* is to identify the effectiveness of Complementary *mind-body-spirit therapies, manipulative and body-based therapies*, and *energy therapies* in children with autism.

**Method:** Literature search method using *ProQuest, PubMed*, and *Science Direct databases* with include the keywords Complementary therapy in children with autism or *Complementary therapy in children with autism*. The search was limited to publication criteria in the 2018-2022 timeframe, full-text articles, and not review articles.

**Results:** Of the study showe that Complementary therapies are effective in improving development in children with autism consisting of *mind-body-spirit therapies* (music, yoga, Qur’an Murrotal Audio) effective in improving: communication and social skills also brain connectivity, the concentration, and attention span, and sleep quality, *manipulative and body based therapies* (physical activity, Tai Chi Chuan, *massage*) are effective in improving social interaction and communication skills, reducing stereotyped behavior, and preventing the risk of autism, *energy therapies* (reflexology, hypnotherapy, and *healing touch*) are effective in reducing the symptoms of autism and constipation and overcome hyperactive behavior.

**Conclusion:** Complementary *mind-body-spirit therapies, manipulative and body-based therapies*, and *energy therapies* are effective in improving the development of children with autism.

## Introduction

Children with Special Needs (ABK) experience abnormalities or anomalies both in terms of physical, mental, intellectual, social, and emotional aspects (Sijabat, 2018). One type of ABK that requires special attention and affects children’s lives is autism because children with autism have complex developmental disorders that significantly affect verbal and nonverbal communication, social interaction, and behavior.

Autism can occur in all conditions, occurs in all countries, races, and all social classes. Based on data published by the World Health Organization (WHO), there is an increase in the number of autism every year in Indonesia. The incidence of autism is increasing globally, including in Indonesia. From 2012 to 2021, the daily increase in autistic children is 147, so in 10 years there will be at least 529,200 cases, so that in 2022 it is estimated that as many as 2.4 million children in Indonesia suffer from autism. Autism is more common in boys, with a prevalence of 1:37, while in girls 1: 151. According to the prevalence data, Indonesia, which has a population of 237.5 million with a population growth rate of 1.14%, is estimated to have a population growth rate of 1.14%. 4 million people with autism, while the WHO predicts 1 in 160 children in the world suffer from autism spectrum disorders. Autism is a complex and lifelong developmental disorder, usually occurring during early childhood and can impact social skills, communication, relationships, and self-regulation (Autism Society of America, 2020). Autism has early signs that parents or caregivers or pediatricians can notice before the child reaches 1 year of age. On average, the symptoms become more consistent when the child is 2 or 3 years old. Some cases suggest that the functional injuries associated with autism may be mild and not noticeable until the child starts school, then their deficiency can be seen when they are among their peers (American Psychiatric Association, 2021)

Interventions that are commonly used to reduce anxiety behavior in children with autism are taking medication. However, this can cause mild to severe side effects, therefore Complementary and alternative therapies are sought by parents for their children with autism (Vidyashree et al., 2019). Complementary therapy is a non-pharmacological treatment that is used as an additional treatment. The use of Complementary therapies in the general population globally has increased significantly in the last 20 years, so it is important for nurses to be familiar with it (Siedlecki, 2021).

The researcher aims to conduct *a literature review* by the topic of Complementary therapy in children with autism in which there are nine types of Complementary therapies, sourced from the results of previous studies and data on children with autism that have been obtained. Researchers also have another reason that underlies researchers to conduct *a literature review*, namely in Indonesia the treatment of children with autism still focuses on providing pharmacological therapy, so that it can reduce the health status and quality of life of children with autism. The results of this *literature review* are expected to be one of the input materials, especially for nursing science in choosing the correct intervention according to the health conditions of children, especially children with autism.

## Search

This research uses a *literature review approach the databases* used are *ProQuest, PubMed*, and *Science Direct*. The strategy used to search the literature by enter the keywords Complementary therapy in children with autism or *Complementary therapy in children with autism*. The inclusion criteria in this research were published in the last 5 years (2018-2022), experimental research in Indonesian and English, full *text*, and with the theme of Complementary therapy in children with autism. Exclusion criteria in this research were published in a span of more than 5 years, articles with the type of *literature review* and *systematic review*, as well as in Indonesian and English forms.

There were 6,389 articles from the *ProQuest database*, 295 articles from *PubMed*, and 2,490 articles from *Science Direct*, so the total articles obtained are 9,174 articles. After *screening* based on inclusion and exclusion criteria, 9 research articles were obtained consisting of 7 international articles and 2 national articles.

**Table 1.**
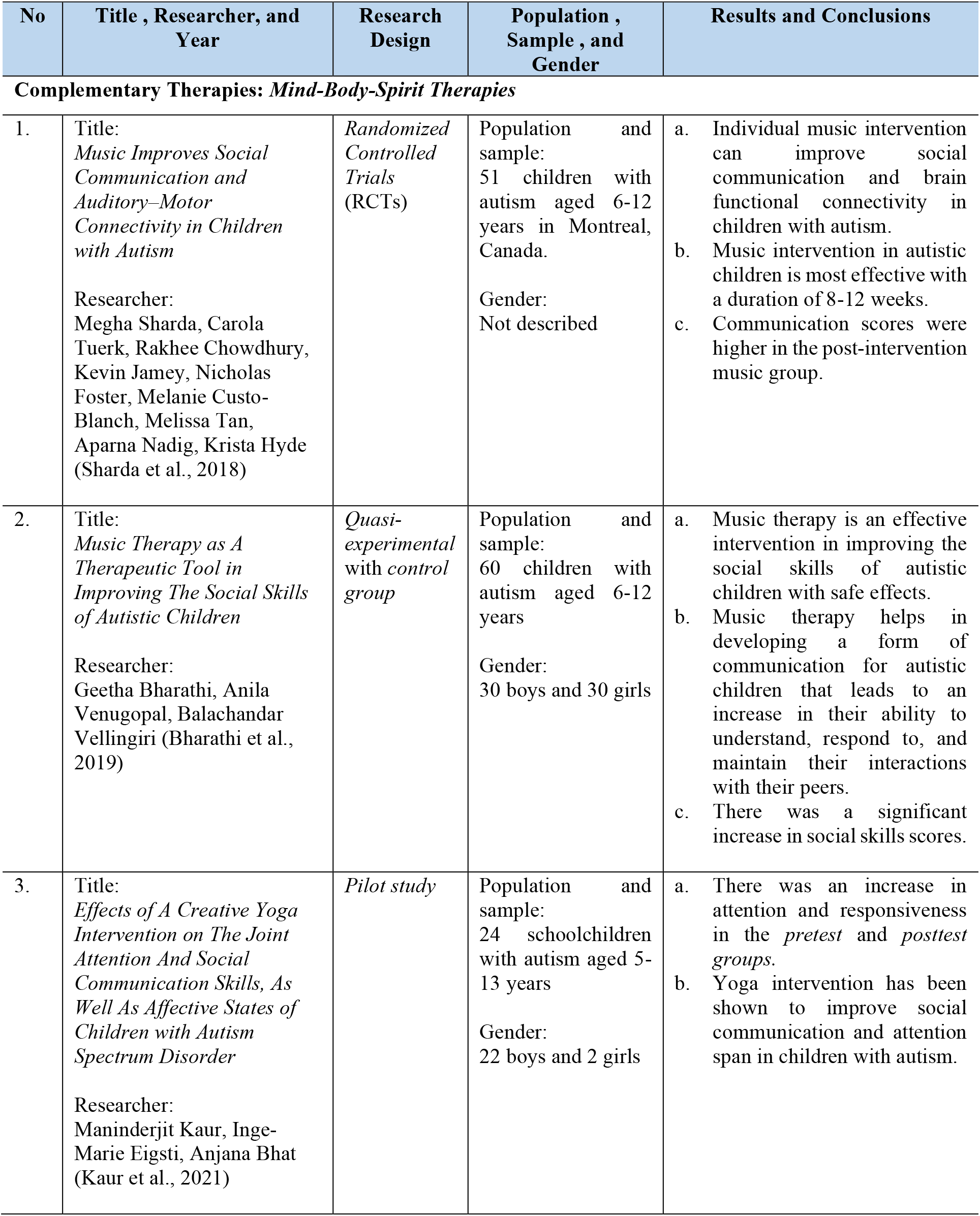
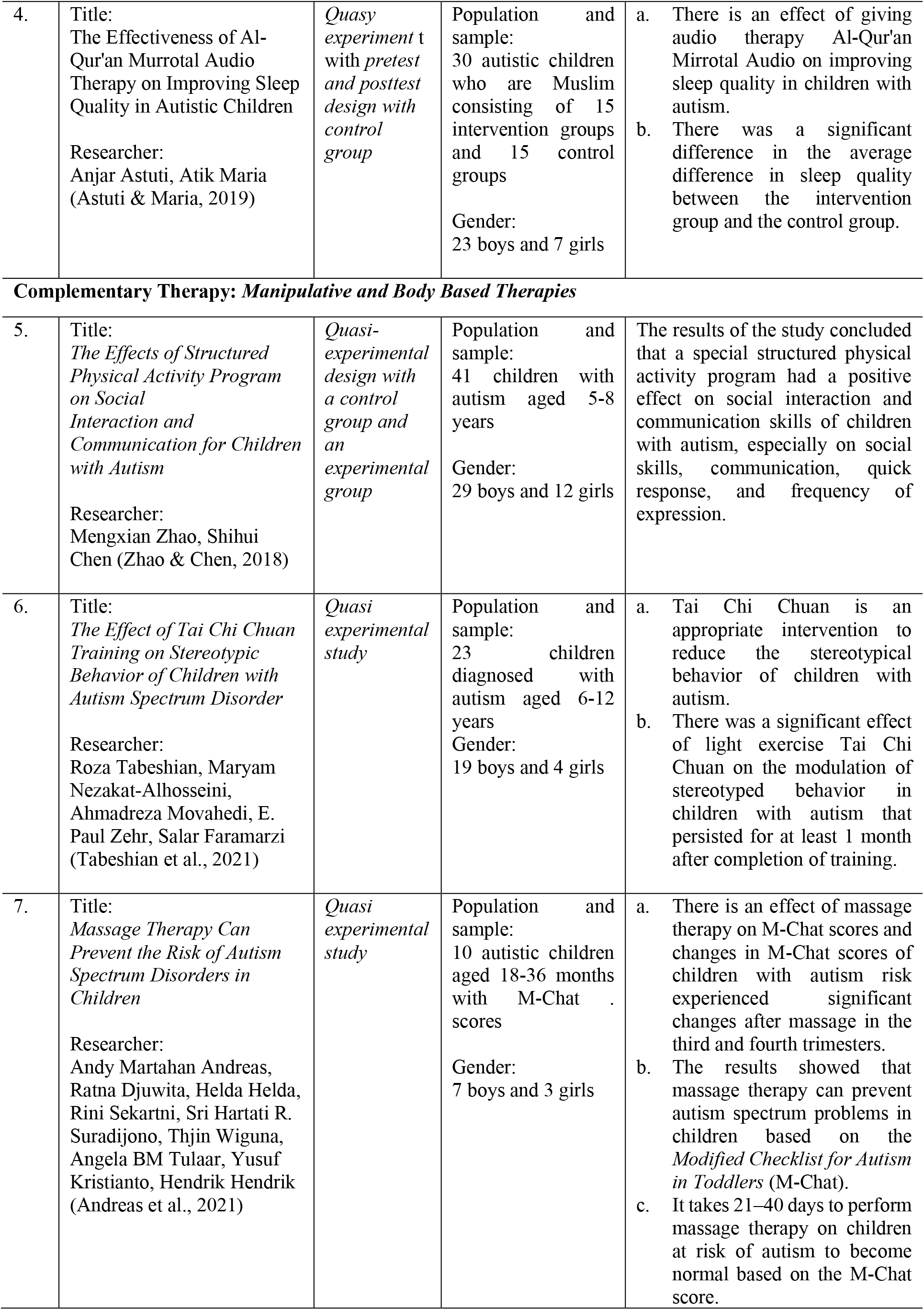
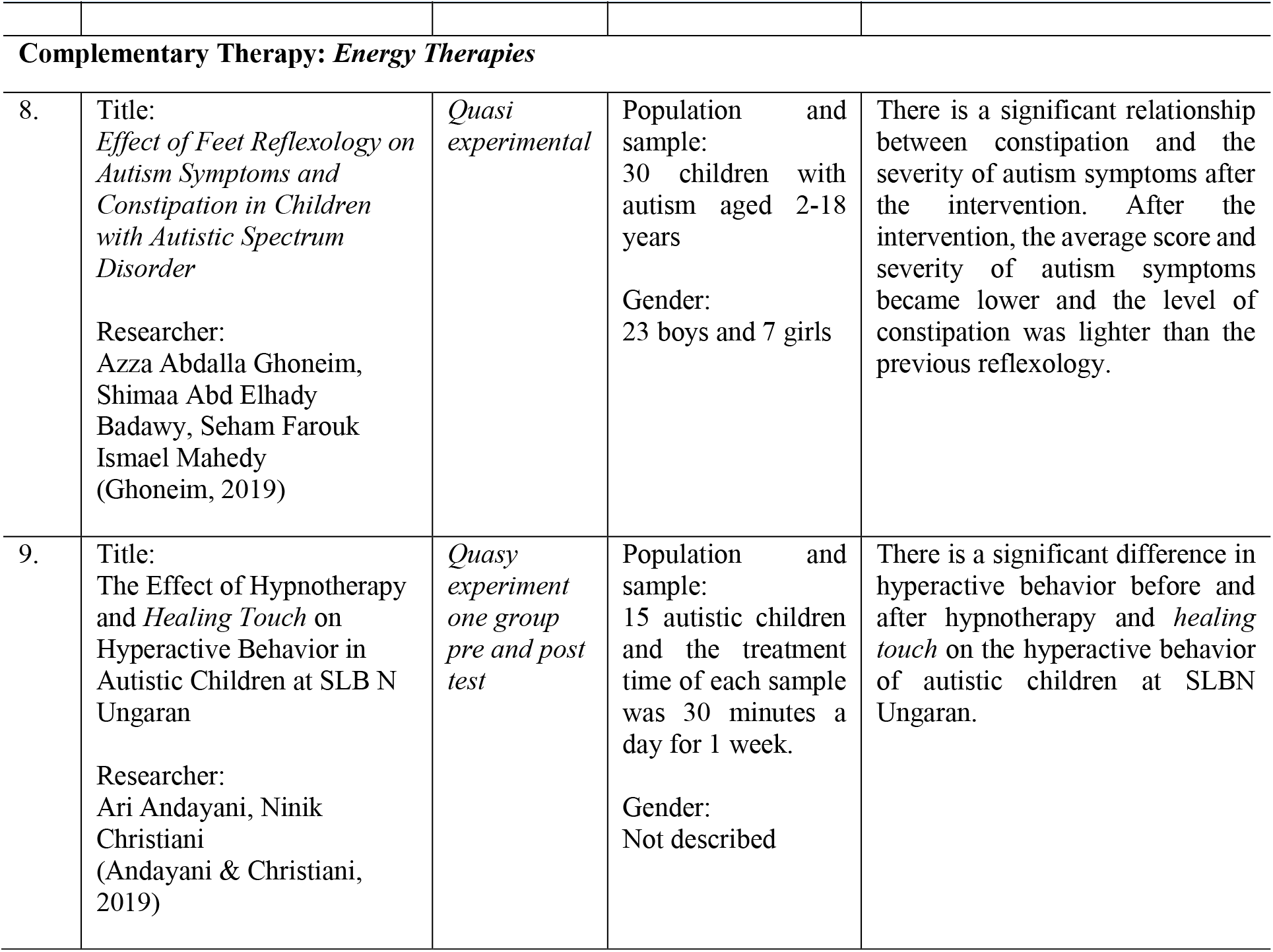
Results of Data Synthesis.

## Results and Discussion

The *literature review research* was conducted on nine articles classified into three based on the type of Complementary therapy, namely *mind-body-spirit therapies* with a total of 4 articles, *manipulative and body-based therapies* in a total of 3 articles, and *energy therapies* with a total of 2 articles. The analysis results of gender in nine articles showed that men were the majority of respondents compared to women, because autism was 4 times more common in men. These findings are in line with research (Rahmawati, 2018) which states that autism is more common in boys than girls with a ratio of 3:1 or 4:1. The reason is hormones, men produce more testosterone and women produce more estrogen (Astuti & Maria, 2019; Wati et al., 2020).

The age range of respondents in nine articles is dominated by school-age children, namely 6-12 years, because at that age the abnormalities that occur in children with autism can be seen clearly, especially in social interaction disorders because the older the child with autism, the more visible the symptoms. And the interventions provided involve the direct participation of children (Kurniawan, 2021; Nadeem et al., 2021).

### 1. Mind-Body-Spirit Therapies for Autistic Children

Research on music as a Complementary therapy for children with autism conducted by (Sharda et al., 2018) and (Bharathi et al., 2019) concluded that music can improve social abilities and skills. These findings are in line with previous research conducted by (LaGasse, 2017) which stated that the use of musical stimulation and musical involvement was useful as a basis for socialization in children with autism which led to an increase in social skills. In the first article, music intervention was carried out for a duration of 8-12 weeks, besides being able to improve social communication, music was also useful in increasing intrinsic connectivity in school-age children (Sharda et al., 2018). In the second article, the provision of music intervention was divided into 2 groups, namely active and passive which was given for 3 months. The musical intervention given to the active group was singing, dancing, and playing musical instruments while listening to music, while the music intervention in the passive group only listened to music alone without interaction. Of the two groups, the active group showed better results compared to the passive group which only received music intervention without further interaction (Bharathi et al., 2019). Children with autism who experience disturbances in social interaction are caused by a lack of ability to regulate the process of social skills which include planning, initiating, and following up with complex motor skills. Rhythm and music can provide unique accommodations for these deficits, because musical stimuli can help plan and execute motor patterns, therefore music interventions have been shown to improve social skills (LaGasse, 2017).

In research on yoga, the definitions (Kaur et al., 2021) show that yoga interventions are useful in improving social communication and attention-focusing in children with autism. The yoga referred to in this research is creative yoga because it consists of the main components of yoga (pose, breathing, and relaxation) made fun and creative using songs, stories, games, also props that are appropriate for the age of the child. Regular yoga practice will have an impact on both branches of the autonomic nervous system by increasing parasympathetic activation and reducing sympathetic nervous system activation of children.

The next research article discusses the Al-Qur’an Murrotal Audio. Children with autism often experience disturbances or problems in sleep patterns which if left unchecked can increase inappropriate behavior such as self-injury, aggression, and tantrums (Astuti & Maria, 2019). Handling sleep quality problems in children with autism is needed so that inappropriate behavior is more controlled (Anam et al., 2019). Audio therapy can be used as an option that is used as an effort to prevent and promote problems with sleep quality. The intervention of listening to the Murrotal sound of the Qur’an is one of the Complementary therapies that is quite effective and better than other audio therapies that are useful as relaxation encouragement, speech therapy, and brain wave therapy, so as to improve sleep quality in children with autism. Therapy is given for 4 days with 2x therapy in 1 day during the day and at night using audio therapy Al-Qur’an Murrotal Audio Surah Ar-Rahman with a pressure of 60 dB for 12 minutes 15 seconds sung by Muhammad Thaha Al-Junayd through laptop media and DVD (Astuti & Maria, 2019). A study that compared brain waves when listening to the recitation of the Qur’an and listening to music based on EEG *(Electroencephalogram) signals* explained that murottal Al-Qur’an therapy can form delta waves in the frontal lobe as the center of intellectual and emotional, including controlling communication skills. And social interaction, as well as the central lobe as a center for controlling movement (Astuti et al., 2017).

### 2. Manipulative And Body Based Therapies for Autistic Children

Children diagnosed with autism show delays in communication and social skills, as well as exhibit repetitive behavioral patterns (Tarr et al., 2020). Research conducted by (Zhao & Chen, 2018) concluded that physical activity programs proved effective in improving skills in social interaction and communication in children with autism. The activity program was carried out for 12 weeks with a directional design and elements of integrated social interaction or communication, every week 2 sessions, one 60-minute session, so that a total of 24 sessions, after 12 weeks showed that the intervention of a structured physical activity program had a positive effect. On communication for children with autism and improve their overall communication such as saying thank you and being able to make eye contact while talking.

Research that has been carried out by researchers (Tabeshian et al., 2021) examines the effects of Tai Chi Chuan exercises on the stereotypical behavior of children with autism. Through this research, Tai Chi Chuan is proven to reduce the stereotypical behavior of children with autism. The intervention was carried out for 45 minutes 3 days a week for 12 weeks which was divided into 3 sessions, namely warm-up for 10 minutes, Tai Chi Chuan exercise for 25 minutes, and cooling down for 10 minutes. During the training session participants were asked to imitate the movements and postures of the trainer. The trainer also provides encouragement and motivation, as well as physical guidance during training. Therapeutic interventions are very useful to reduce stereotyped behavior is very important to improve and support development, especially children with autism. Tai Chi Chuan exercise can reduce the activity of the sympathetic nervous system, increase parasympathetic activity, and stabilize serotonin levels because in children with autism there is dysfunction of parasympathetic tone regulation. Tai Chi Chuan exercises can also change brain morphology and stabilize motor control in children with autism because of the balanced coordination of accurate limb movements combined with complex and integrated whole body movements.

Complementary therapy carried out by researchers (Andreas et al., 2021) namely *massage therapy* (massage) has been proven to prevent autism problems in children based on the Modified Checklist for Autism in Toddler (M-Chat). Massage is often used as a Complementary therapy in the treatment of children with autism, as many as 11%-16% of people with autism undergo massage therapy (Ruan et al., 2021). To achieve maximum results, it takes 21–40 days to perform massage therapy on children at risk of autism until they become normal based on the M-Chat score. Children are given massage therapy for 40 days which is divided into four periods, where each period takes 10 days. The measurement of the M-Chat score was carried out before and after the intervention was given to determine the level of autism risk.

### 3. Energy Therapies for Autistic Children

Research by (Ghoneim, 2019) discusses the effects of reflexology on symptoms of autism and constipation in children with autism. Reflexology is an ancient therapeutic art that combines holistic pressure and invasive touch, applying precise hand and finger pressure to specific points on the feet called reflex sites. These reflex sites are believed to be associated with the internal organs of the body. The intervention was carried out once a week for 6 weeks with a time of 30 minutes (15 minutes per leg). The implementation is divided into 3 stages, namely preparation, work, and evaluation. At the end of the sixth week, a reassessment of the symptoms of autism and constipation was carried out. The results of the research concluded that there was a decrease in autism symptom severity scores and a lower level of constipation. Reflexology is a simple procedure that is useful and gives positive results in constipation in children with autism (Ghoneim, 2019). This statement is in accordance with research that has been carried out by (Azari et al., 2021) which states that reflexology has been proven to be effective in improving constipation symptoms. Reflexology is influential in regulating the balance of the autonomic nerves and increasing the blood supply to the projected organs.

Research on the effect of hypnotherapy and *healing touch* on hyperactive behavior in children with autism studied by researchers (Andayani & Christiani, 2019) concluded that hypnotherapy and *healing touch* can overcome hyperactive behavior and increase compliance in children with autism. There was a decrease in the level of hyperactivity during the intervention when the first 10 minutes the child was able to receive hypnotherapy and *healing touch* and tried to focus his attention, in the next 10 minutes the child had become calmer and more organized, and in the last 10 minutes to 15 minutes after the intervention the child became calm. From the research results of respondents who have done hypnotherapy and *healing touch*, it is found that there is a decrease in the hyperactive behavior of children with autism before and after being given hypnotherapy and *healing touch* for 30 minutes.

## Conclusion

The conclusion that can be drawn after analyzing nine articles is that Complementary therapies are effective in improving development in children with autism consisting of 4 articles on *mind-body-spirit therapies*, 3 articles on *manipulative and body-based therapies*, and 2 articles on *energy therapies*. Complementary *mind-body-spirit therapies* effective in improving development in children with autism which includes music can improve communication and social skills, yoga is useful in improving social skills and concentration of attention, as well as Al-Qur’an Murrotal Audio can improve sleep quality. Complementary *manipulative and body based therapies are* effective in improving development in children with autism which include physical activity useful in improving interaction and social skills, Tai Chi Chuan can reduce stereotyped behavior, and massage is useful for preventing the risk of autism. Complementary *energy therapies* are effective in improving development in children with autism which include reflexology that can reduce symptoms of autism and constipation, and hypnotherapy and *healing touch* are useful in overcoming hyperactive behavior.

## Data Availability

All data produced in the present work are contained in the manuscript

